# Sex Disparities in Bladder Cancer Diagnosis: Evidence from the UK Biobank

**DOI:** 10.1101/2025.06.04.25328951

**Authors:** Dorit Shweiki

## Abstract

A sex-stratified analysis of 4,848 bladder cancer cases and matched controls from the UK Biobank identified hematuria as the strongest predictor across all groups. Urinary tract infections were predictive in all female age groups—most strongly under age 50—and in younger males only. These findings highlight age- and sex-based diagnostic disparities, with delayed recognition more likely in women, underscoring the need for improved clinical awareness and response.

In this study, we looked at how symptoms like blood in the urine and urinary tract infections predict bladder cancer. We found that urinary infections can signal bladder cancer in women and younger men, but may be overlooked. Improving symptom recognition could lead to earlier diagnosis, especially for women.

## Introduction

Bladder cancer (BC) is the fourth most common cancer among men, with incidence and mortality rates approximately four times higher in men (9.5 and 3.3 per 100,000, respectively) than in women (2.4 and 0.9, respectively). Its occurrence varies significantly worldwide, with the highest rates for both men and women in Western and Southern Europe and North America, and the lowest for men in Middle Africa, South Central Asia, and Western Africa [1, 2]. BC development is linked to advanced age, male sex, cigarette smoking, and exposure to occupational and environmental toxins[1].

Hematuria is the most common presenting symptom of BC [3]. In adults, any episode of hematuria—microscopic or gross—carries a substantial risk of underlying malignancy [4]. Its predictive value increases with patient age and is higher among men than women [3]. Moreover, the type of hematuria may reflect disease stage, as gross hematuria presentation has been associated with a higher likelihood of muscle-invasive disease and high-grade histology [5].

Second to hematuria, urinary tract infections (UTIs) are often documented in BC patients, and this connection has been widely investigated [6]. A comprehensive meta-analysis concluded that a history of chronic UTIs significantly increases BC risk [7]. An earlier study in nearly 3,000 BC cases found that having three or more UTIs was linked to a twofold increase in BC risk, particularly for squamous cell carcinoma [8]. However, conflicting findings exist regarding the association between multiple UTIs and BC risk. Most research indicates a significant increase in BC risk with multiple UTIs [6, 8, 9], yet a study by Jiang and colleagues demonstrated a markedly reduced risk of BC in patients with recurrent UTIs, potentially explained by antimicrobial treatment [10]. Moreover, a meta-analysis by Bayne et al. questioned causality, attributing heterogeneous results to non-comparable or inaccurate UTI data, differences in geographic cohorts, and the influence of publication year on study accuracy [11].

Early cancer symptoms can overlap with those of more common conditions, often causing diagnostic delays. For instance, pancreatic cancer may present with hyperglycemia and be erroneously diagnosed as new-onset type II diabetes [12]. Ovarian cancer often mimics irritable bowel syndrome or UTIs, while lung cancer may be confused with chronic bronchitis or pneumonia [13, 14]. These challenges arise because early-stage cancers tend to have nonspecific or overlapping symptoms easily mistaken for benign conditions. In addition, physicians typically prioritize ruling out common disorders first, and the absence of robust early screening methods for some cancers can further delay accurate diagnosis.

Indeed, a similar diagnostic challenge occurs when evaluating BC symptoms. In some instances, what appears to be a recurrent UTI is BC provoking chronic bladder irritation or harboring bacteria. Consequently, patients with undiagnosed BC may experience recurrent “UTI-like” symptoms for an extended period before receiving the correct diagnosis. This overlap in presentation often leads to misdiagnosis or delayed diagnosis, especially among women.

A similar pattern emerges with hematuria. Studies indicate that hematuria is often under-investigated in clinical practice, and only a relatively small proportion of individuals with new-onset hematuria are referred to a urologist [4].

Sex disparities in BC diagnosis, specifically regarding UTIs and hematuria, are particularly concerning. Evidence suggests that women with BC are more likely than men to receive an initial misdiagnosis of UTI. In women, hematuria may be attributed to infection or menstruation rather than cancer, often leading to delayed or incorrect diagnoses. As a result, women tend to be diagnosed at a later stage. Certain studies indicate that the first diagnosis of muscle-invasive BC occurs nearly twice as often in women as in men [15].

Considering the higher rate of BC misdiagnosis among women and the inconclusive evidence regarding UTIs as BC predictors, this brief communication examines sex disparities in UTI occurrences and hematuria among BC patients, and their correlation to disease stage.

## Methods

### Data Source and Study Cohort

This retrospective case–control study used data from the UK Biobank (UKB), a population-based prospective cohort of ∼502 000 adults aged 40– 69 years at enrolment between 2006 and 2010. We analyzed the linked hospital inpatient data (Hospital Episode Statistics; HES), national cancer registry and national death registry extracts that were downloaded on 26 April 2023 under UKB application 89496. The analytic dataset comprised 4,848 bladder-cancer cases (3,630 men and 1,218 women) and 14,544 age-and sex-matched controls, yielding a case-to-control ratio of 1: 3. All participants provided written informed consent; the study operates under UK Biobank’s generic ethical approval (REC 11/NW/0382) [16].

### Inclusion Criteria

Eligible participants had complete linkage to hospital inpatient, cancer-registry and death-registry records with valid entries for sex, year of birth and baseline assessment. Cases were participants with at least one ICD-10 C67.x code (malignant neoplasm of bladder) recorded in any of the linked secondary-care sources; the earliest such record defined the index date. Controls were participants without C67.x codes in any source up to the download date (26 April 2023), alive on their assigned pseudo-index date, and free of other malignant neoplasms of the urinary tract (ICD-10 C65–C66, C68).

### Exclusion Criteria

Participants were excluded if they had withdrawn consent; lacked a valid self-reported sex value; presented implausible year-of-birth values or missing baseline assessment dates that precluded accurate age calculation; in the control cohort, had any invasive cancer diagnosis (ICD-10 C00–C97, non-melanoma skin cancer excepted) recorded before baseline; had a bladder-cancer code documented only on a death certificate without an accompanying diagnosis date; or had duplicate hospital episodes for which the earliest distinct date was retained.

### Index Date and Control Matching

The index date for each case was the first record of bladder-cancer (C67.x) diagnosis. Three controls were frequency-matched on sex and year of birth (± 1 year) to each case (overall 1: 3 ratio) and were assigned the same index date as their corresponding case.

### Exposure and Covariate Assessment

Linked cancer-registry, HES (primary and secondary diagnoses) and death-registry records were searched for diagnoses in the 10 years preceding the index date. Hematuria was defined by ICD-10 R31 or any code beginning with N02, while urinary-tract infection (UTI) was defined by ICD-10 N39.0.

Participants were classified as exposed if ≥ 1 qualifying diagnosis occurred in that interval. Although event dates and counts were retained, only this binary exposure indicator entered the regression models.

Comorbidities - irritable bowel syndrome, hypertension, asthma, diabetes mellitus and ischemic heart disease—were coded as present if recorded in the hospital inpatient datasets (primary or secondary diagnosis fields) or the cancer registry at any time before the index date. Smoking status was derived from baseline questionnaires and categorized as never (reference), former or current smoker.

### Statistical Analysis

Associations between clinical or demographic predictors and bladder-cancer status were quantified using Firth logistic regression, chosen for its robustness against bias and instability when events or predictors are rare; the method mitigates small-sample bias and separation. Results are reported as odds ratios (ORs) with 95 % confidence intervals; two-sided p < 0.05 was considered statistically significant.

### Stratified analysis by sex and age

Separate models were fitted for men and women and for three age groups (< 50, 50–64, ≥ 65 years). All predefined strata contained ≥ 20 participants and at least one outcome event.

### Bladder-cancer-only analysis

Within cases, Firth logistic regression models were fitted **separately for men and women** to evaluate predictors of tumor invasiveness and all-cause mortality. Invasiveness was classified via tumor-behavior codes (invasive vs non-invasive); mortality status was taken from death-registry linkage.

### Computational Environment and Software

All analyses were conducted in Python 3.10.12; data cleaning and transformation were performed with pandas 2.2.3, and penalized logistic models were fit with the firthlogist package (v 0.5.0) within a dedicated virtual environment. No alternative model-selection procedures were employed; confidence intervals were those returned by the package.

This design enabled examination of both risk factors for incident bladder cancer and determinants of disease aggressiveness, while explicitly controlling for sex-and age-related heterogeneity.

## Results

A UK Biobank cohort of 4,848 BC patient records (3,630 males and 1,218 females), and age- and sex-matched controls were analyzed. We retrieved information on BC diagnosis, tumor type and invasiveness, disease outcome, and the number of UTI and hematuria events in the ten years prior to BC diagnosis (and age-related index for controls). Data were stratified by sex, and analyses were conducted separately for males and females to avoid masking effect. Smoking status and comorbidities were assessed for their contribution to BC risk, while associations between UTIs, hematuria events, and BC invasiveness or outcomes were evaluated for BC patients.

We assessed the main predictors of BC risk by sex and age using Firth logistic regression. Hematuria in the ten years preceding BC diagnosis was the strongest and most consistent predictor across all sex and age groups. Odds ratios for hematuria ranged from 5.42 in older males (65+) to 56.32 in females under 50 (all p < 0.0001), reflecting its central role as an early clinical indicator. UTI events were also predictive, particularly in younger subgroups: females under 50 showed an odds ratio of 22.16 (p = 0.0003), and males under 50 had an odds ratio of 8.09 (p = 0.0165). In contrast, the predictive value of UTIs diminished with age, becoming statistically nonsignificant in older groups. The magnitude of these odds ratios suggests that hematuria—and to a lesser extent, UTIs—likely represent early manifestations of undiagnosed bladder cancer rather than causal risk factors. Smoking, both past and current, was associated with increased BC risk, particularly in older subgroups. Comorbidities such as hypertension and diabetes showed significant associations, primarily in middle to older age groups (Table S1).

We next focused solely on BC patients to evaluate whether hematuria and UTI events were associated with tumor invasiveness and mortality. Hematuria was inversely associated with invasive disease in two female subgroups (<50 and 65+) and in males aged 50–64, potentially reflecting earlier detection due to symptom onset or characteristics of non-invasive tumors. UTI was not predictive of tumor invasiveness in any age or sex group. However, in patients aged 65 and older—both males and females—UTIs were associated with increased mortality. Mortality was primarily associated with comorbidities, particularly diabetes and heart disease, and with smoking in older male patients (Table S2).

**Table 1.**
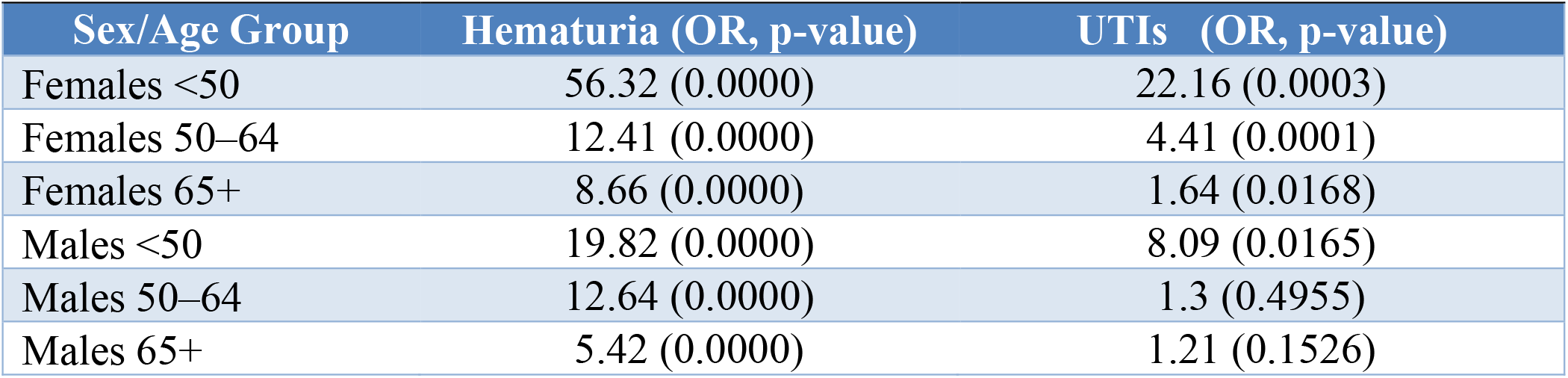
Hematuria and UTI Predictors of Bladder Cancer. Odds ratios and p-values for hematuria and urinary tract infections in the ten years preceding diagnosis as predictors of bladder cancer across sex-by-age subgroups.

## Discussion

These findings reinforce hematuria’s established role in BC diagnosis and highlight its importance as a critical screening indicator for both sexes. More notably, the results underscore UTI as a significant predictive factor, particularly among younger female patients. Given the higher prevalence of UTIs in women, physicians may be inclined to attribute such symptoms to benign infections, potentially leading to delayed or missed BC diagnoses. Our results call for heightened clinical awareness of sex-and age-specific patterns in UTI and hematuria presentations. Rather than dismissing UTIs in women as routine, clinicians should consider them potential red flags, especially in younger women, where earlier recognition may expedite BC diagnosis and improve outcomes.

Limitations of this study include reliance on routinely collected diagnostic codes, which may undercapture or misclassify UTI and hematuria events, particularly in primary care settings not linked to the UK Biobank. Additionally, the UK Biobank cohort may not fully represent the general population due to healthy volunteer bias, which could potentially limit its generalizability.

## Conclusions

This study reveals a previously underrecognized sex disparity in bladder cancer diagnosis: while hematuria remains a consistent predictor across all groups, urinary tract infections (UTIs) emerged as strong predictors of bladder cancer only in women and younger men. These findings suggest that in specific populations, particularly younger women, UTIs may reflect early malignant processes rather than benign infections. The diagnostic significance of UTIs is often underappreciated in women, potentially contributing to delayed detection and poorer outcomes. Our results support the need for sex- and age-specific symptom interpretation in bladder cancer risk assessment and clinical decision-making.

## Data Availability

This research was conducted using the UK Biobank Resource under approved project number 89496. Data are available from the UK Biobank but are not publicly available due to data access restrictions

https://www.ukbiobank.ac.uk/

## Contributors

D.S. conceived the study, conducted data analysis, interpreted the results, and wrote the manuscript.

## Acknowledgments

This research has been conducted using the UK Biobank Resource under Application Number 89496.

## Data Sharing

This research was conducted using the UK Biobank Resource under approved project number 89496. Data are available from UK Biobank but are not publicly available due to data access restrictions.

Supplementary Tables: Tables S1 and S2

