## Supplementary material for "Sex Disparities in Bladder Cancer Diagnosis: Evidence from the UK Biobank": Tables S1 and S2

**Table S1**: Predictors Of Bladder Cancer

| Predictor | Measure | Females <50 | Females 50–64 | Females 65+ | Males <50 | Males 50–64 | Males 65+ |
|---|---|---|---|---|---|---|---|
| **Hematuria** | **Odds Ratio** | 56.32 | 12.41 | 8.66 | 19.82 | 12.64 | 5.42 |
| | **P-value** | 0 | 0 | 0 | 0 | 0 | 0 |
| **UTI** | **Odds Ratio** | 22.16 | 4.41 | 1.64 | 8.09 | 1.3 | 1.21 |
| | **P-value** | 0.0003 | 0.0001 | 0.0168 | 0.0165 | 0.4955 | 0.1526 |
| **Hypertension** | **Odds Ratio** | 1.3 | 1.34 | 1.43 | 1.96 | 1.55 | 1.88 |
| | **P-value** | 0.4873 | 0.024 | 0.0003 | 0.0042 | 0 | 0 |
| **Diabetes** | **Odds Ratio** | 1.62 | 1.54 | 1.1 | 1.05 | 1.32 | 1.17 |
| | **P-value** | 0.382 | 0.02 | 0.5019 | 0.8716 | 0.0035 | 0.0198 |
| **IBS** | **Odds Ratio** | 0.62 | 1.27 | 1.61 | 1.89 | 1.81 | 1.25 |
| | **P-value** | 0.6254 | 0.3063 | 0.0235 | 0.3154 | 0.0111 | 0.1993 |
| **Asthma** | **Odds Ratio** | 2.07 | 1.38 | 1.05 | 2.2 | 1.19 | 1.02 |
| | **P-value** | 0.0947 | 0.0629 | 0.7039 | 0.0179 | 0.1267 | 0.8123 |
| **Heart** | **Odds Ratio** | 2.73 | 1.17 | 1.07 | 1.21 | 1.11 | 1.05 |
| | **P-value** | 0.0217 | 0.3008 | 0.5555 | 0.4201 | 0.2074 | 0.4053 |
| **Smoking Former** | **Odds Ratio** | 2.58 | 1.29 | 1.4 | 1.26 | 1.85 | 1.56 |
| | **P-value** | 0.0064 | 0.0447 | 0.0005 | 0.3193 | 0 | 0 |
| **Smoking Current** | **Odds Ratio** | 1.78 | 2.27 | 2.37 | 1.89 | 2.46 | 2.27 |
| | **P-value** | 0.1928 | 0 | 0 | 0.018 | 0 | 0 |

**Table S1.** Odds ratios and p-values for clinical and demographic predictors of bladder cancer across sex and age-stratified subgroups. Odds ratios were derived from Firth logistic regression models. Smoking status was modeled as a categorical variable with never smokers as the reference category and included separate indicators for former and current smokers.

**Table S2**: Predictors of Invasiveness and Mortality

| Predictor | Outcome | Female <50 | Female 50-64 | Female 65+ | Male <50 | Male 50-64 | Male 65+ |
|---|---|---|---|---|---|---|---|
| Urinary Tract Infection | Invasiveness | 1.32 (0.6665) | 0.73 (0.5072) | 1.23 (0.5544) | 0.56 (0.4798) | 1.05 (0.8947) | 1.04 (0.8654) |
| | Mortality | 1.32 (0.7355) | 0.88 (0.8098) | 2.47 (0.0033) | 0.7 (0.7602) | 1.25 (0.6258) | 2.21 (0.0000) |
| Hematuria | Invasiveness | 0.2 (0.0335) | 0.65 (0.0891) | 0.47 (0.0014) | 0.47 (0.1037) | 0.73 (0.0494) | 1.05 (0.6912) |
| | Mortality | 0.29 (0.2981) | 0.56 (0.0622) | 1.01 (0.9446) | 0.61 (0.3973) | 0.88 (0.4489) | 1.35 (0.0098) |
| Irritable Bowel Syndrome | Invasiveness | 0.08 (0.0723) | 0.79 (0.5096) | 1.36 (0.4242) | 1.81 (0.6762) | 0.84 (0.6184) | 1.08 (0.8137) |
| | Mortality | 2.64 (0.5849) | 0.48 (0.0949) | 0.71 (0.3602) | 1.11 (0.8340) | 0.53 (0.1268) | 0.76 (0.3658) |
| Hypertension | Invasiveness | 3.43 (0.0630) | 1.03 (0.9005) | 0.89 (0.5421) | 2.13 (0.0962) | 0.75 (0.0564) | 1.0 (0.9506) |
| | Mortality | 0.96 (0.6676) | 1.3 (0.2820) | 0.72 (0.1065) | 2.41 (0.0574) | 0.93 (0.6321) | 1.0 (0.9714) |
| Asthma | Invasiveness | 3.3 (0.1088) | 0.9 (0.6986) | 1.25 (0.4007) | 1.12 (0.8177) | 1.38 (0.1225) | 1.13 (0.4762) |
| | Mortality | 0.25 (0.2205) | 0.97 (0.8949) | 2.06 (0.0027) | 0.85 (0.7885) | 0.96 (0.8374) | 1.34 (0.0475) |
| Diabetes | Invasiveness | 0.21 (0.1107) | 0.9 (0.7307) | 1.1 (0.7198) | 1.58 (0.5208) | 0.93 (0.6673) | 0.97 (0.8166) |
| | Mortality | 0.27 (0.3649) | 2.53 (0.0018) | 1.86 (0.0143) | 2.24 (0.1530) | 1.61 (0.0031) | 1.41 (0.0022) |
| Heart Disease | Invasiveness | 1.22 (0.7847) | 1.43 (0.1568) | 1.32 (0.1872) | 0.54 (0.2059) | 1.08 (0.5920) | 1.07 (0.5084) |
| | Mortality | 1.54 (0.5997) | 1.48 (0.1359) | 1.98 (0.0007) | 1.02 (0.8705) | 1.39 (0.0235) | 1.53 (0.0000) |
| Smoking (Former) | Invasiveness | 1.78 (0.3436) | 1.25 (0.3382) | 0.85 (0.4201) | 2.69 (0.0427) | 1.0 (0.9609) | 0.97 (0.7699) |
| | Mortality | 0.71 (0.6497) | 1.52 (0.1031) | 1.41 (0.0847) | 1.53 (0.4212) | 1.42 (0.0294) | 1.29 (0.0237) |
| Smoking (Current) | Invasiveness | 1.49 (0.5940) | 0.74 (0.2743) | 0.53 (0.0265) | 1.3 (0.5671) | 0.9 (0.5802) | 1.11 (0.5195) |
| | Mortality | 0.57 (0.4986) | 1.64 (0.1111) | 1.48 (0.1806) | 1.94 (0.2192) | 1.88 (0.0016) | 2.0 (0.0000) |

**Table S2.** Odds ratios (OR) and p-values for predictors of tumor invasiveness and mortality among bladder cancer patients, stratified by sex and age group. Each predictor is evaluated separately for its association with either invasiveness or mortality using Firth logistic regression. Odds ratios were derived by exponentiating model coefficients. P-values are based on penalized likelihood ratio tests. Smoking status was modeled categorically, with never smokers as the reference group; "Smoking (Former)" and "Smoking (Current)" reflect separate indicator variables. Statistically significant results ($p < 0.05$) are highlighted in red font.
